# Meta-analysis of Sequencing Data Identifies Candidate Endometrial Cancer Susceptibility Genes

**DOI:** 10.64898/2026.11.23.26368185

**Authors:** Michelle Laker, Tracy A. O’Mara, Amanda B. Spurdle, Marc Tischkowitz, Paul D.P. Pharoah, Emma J. Davidson, Douglas F. Easton, Antonis C. Antoniou, Xin Yang

## Abstract

**Background:** Endometrial cancer is the most common gynaecological malignancy in high-income countries, yet its genetic architecture remains incompletely understood. While pathogenic variants in mismatch repair genes are well-established contributors, the role of rare variants in other genes has not been systematically characterized.

**Methods:** We evaluated associations between rare genetic variants and endometrial cancer risk based on 2,675 cases and 160,654 controls from the UK Biobank and the 100,000 Genomes Project. Gene-level associations were assessed using burden tests, with protein-truncating variants (PTVs) and missense variants (MSVs) predicted to be deleterious analysed separately.

**Results:** Endometrial cancer (EC) risk associations were observed for three established EC susceptibility genes in the rare PTV analysis: *MSH6* (RR = 36.6, 95% CI 25.9–51.8), *MLH1* (RR = 52.5, 95% CI 21.3–129.4) and *MSH2* (RR = 13.0, 95% CI 6.0–28.2). In the rare MSV analysis, *PTEN* was also significantly associated with EC risk (RR = 9.7, 95% CI 4.0–23.7). Beyond these established susceptibility genes, exome-wide significance was observed for 19 additional genes in the PTV analysis, including *PHKG2* (RR = 29.8, 95% CI 9.6–92.8), and eight additional genes in the MSV analysis.

**Conclusion:** This study identifies novel candidate genes associated with endometrial cancer risk, providing a foundation for future replication studies.

## INTRODUCTION

Endometrial cancer is the sixth most diagnosed cancer in women, and accounts for 1% of cancer-related deaths worldwide ^1^. The global incidence rose by 0.69% per year between 1990 and 2019 ^2^, and is projected to increase further due to rising obesity rates and an ageing population ^3^.

Despite its growing burden, susceptibility to endometrial cancer remains under-researched relative to other cancers of similar impact. Epidemiological studies have shown that a family history of endometrial cancer in at least one first- or second-degree relative is associated with an increased risk of endometrial cancer (HR=3.66, 95% CI: 2.63–5.08). ^4,5^. However, to date, only five genes have been established as endometrial cancer susceptibility genes: the four DNA mismatch repair (MMR) genes associated with Lynch syndrome (*MLH1, MSH2, MSH6*, and *PMS2*), and *PTEN* that is associated with PTEN hamartoma tumor syndrome (PHTS) ^4,6,7^.

Germline variants in several other genes, including *POLE, POLD1, MUTYH, FH* and *BRCA1/2*, have been proposed as associated with endometrial cancer risk ^8–11^. However, evidence of association either remains inconsistent, or lacks robust risk estimates ^12^. Beyond rare coding genetic variants, genome-wide association studies (GWAS) have identified multiple common low-risk variants associated with endometrial cancer risk ^13^. Collectively the identified genetic variants explain a small proportion of the endometrial cancer familial risk and suggest that other genetic susceptibility variants remain to be identified.

Here, we performed a meta-analysis combining exome sequencing data from the UK Biobank (UKB) and the 100,000 Genomes (100KG) project to identify candidate susceptibility genes associated with endometrial cancer.

## RESULTS

After filtering, the analysis included 1,956 cases (752 prevalent, 1204 incident) and 148,948 controls from UKB, and 719 cases and 11,706 controls from 100KG. Age distributions are shown in Supplementary Table 1. Burden analyses were conducted for 15,247 genes selected based on rare PTVs, 16,821 genes selected for a combination of rare PTVs and splice-altering genes, and 17,535 genes selected based on rare predicted deleterious MSVs.

### Protein-truncating variants

When carriers were selected for the presence of rare PTVs only, a combined meta-analysis of the UKB and 100KG identified 22 genes exceeding the exome-wide significance threshold, with relative risk (RR) ratios ranging from 4.2 to 66.7 (Figure 1, Table 1). The strongest associations were observed for *MSH6* (RR = 36.6, 95% CI 25.9–51.8), *MLH1* (RR = 52.5, 95% CI 21.3–129.4), and *MSH2* (RR = 13.0, 95% CI 6.0–28.2). An additional 48 genes showed associations at p < 0.0001 (Supplementary Table 2). There was also no observed heterogeneity between the two studies for each of the most strongly associated genes (Supplementary Table 2).

**Figure 1.**
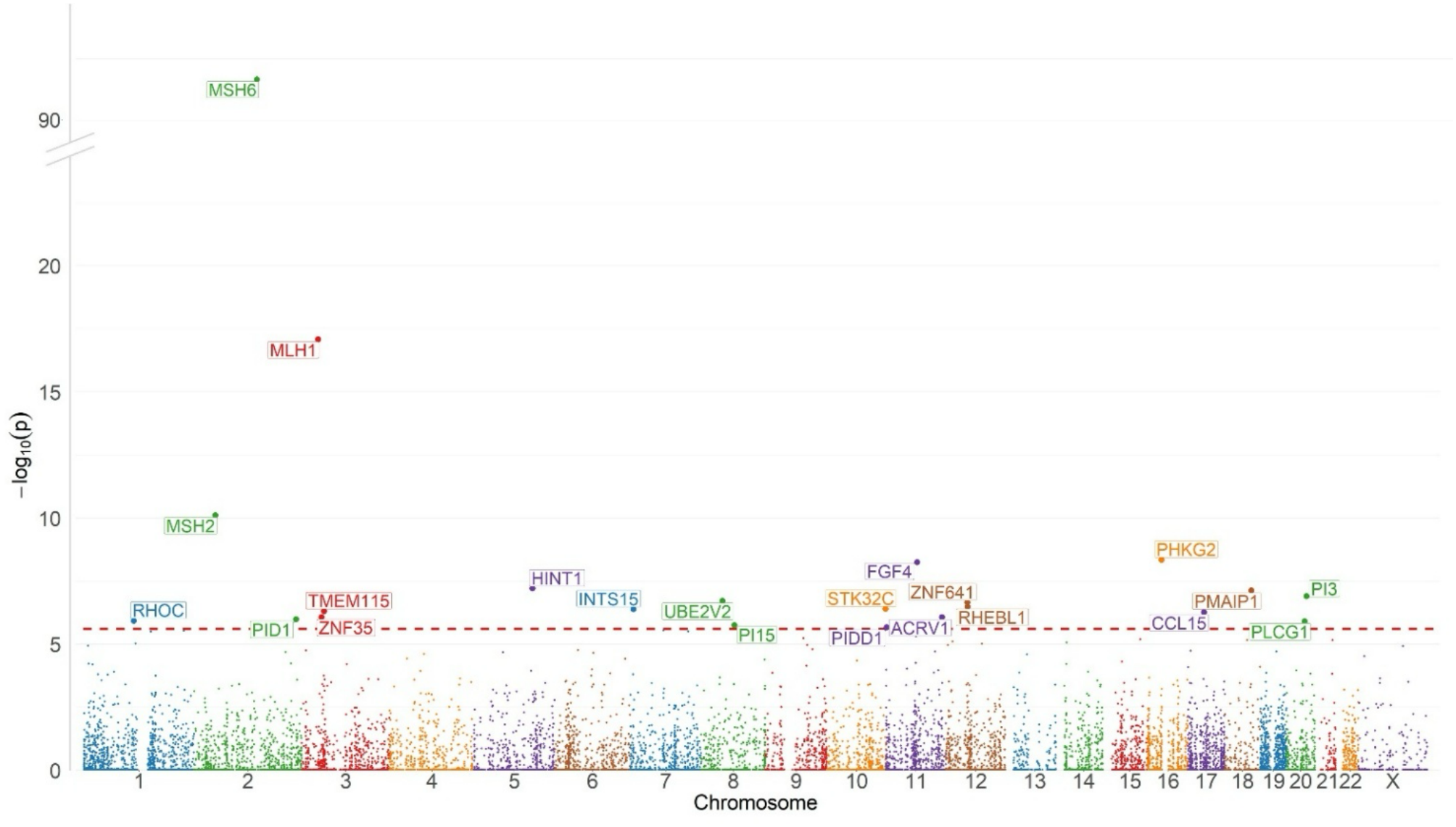
Manhattan plot showing the associations between endometrial cancer and rare protein truncating variants (PTVs) by gene, based on a meta-analysis combining whole-exome sequencing analysis from the UK Biobank with whole genome sequencing data from 100,000 Genomes Project using logistic regression. The red dashed line represents the exome-wide significance threshold of p=2.5×10^−6^.

**Table 1.** Results of the analysis of rare protein-truncating variants (PTVs) in genes associated with endometrial cancer at exome-wide significance (p < 2.5 × 10^−6^) in the meta-analysis of UK Biobank (UKB) and 100,000 Genomes (100KG) Project datasets using logistic regression. The table shows PTV carrier counts among cases and controls, together with relative risks (RRs) and 95% confidence intervals (CIs) from the UKB prevalent and incident meta-analyses, the 100KG analysis, and the meta-analyses combining UKB and 100KG.

| **Gene** | **UKB Counts (Prevalent Cases/ Controls)** | **UKB Counts (Incident Cases/ Controls)** | **UKB Meta Analysis (Prevalent and Incident Cohorts)** |  | **100KG Counts (Cases/ Controls)** | **100KG Analysis** |  | **Meta-analysis (UKB and 100KG)** |  |
|---|---|---|---|---|---|---|---|---|---|
|  |  |  | **Relative Risk (95% CI)** | **p value** |  | **Odds Ratio (95% CI)** | **p value** | **Relative Risk (95% CI)** | **p value** |
| *MSH6* | 16/63 | 17/63 | 38.0 (26.4 - 54.5) | 5.0×10<sup>-86</sup> | 7/5 | 25.3 (7.9 - 81.0) | 5.1×10<sup>-8</sup> | 36.6 (25.9 - 51.8) | 2.2x10<sup>-92</sup> |
| *MLH1* | 4/9 | 2/9 | 52.5 (21.3 - 129.4) | 8.1×10<sup>-18</sup> | 0/1 | - | 0.96 | 52.5 (21.3 - 129.4) | 8.1x10<sup>-18</sup> |
| *MSH2* | 4/38 | 1/38 | 14.4 (5.8 - 36.0) | 1.1×10<sup>-8</sup> | 3/5 | 10.0 (2.4 - 42.4) | 1.7×10<sup>-3</sup> | 13.0 (6.0 - 28.2) | 7.6x10<sup>-11</sup> |
| *PHKG2* | 0/11 | 3/11 | 29.8 (9.6 - 92.8) | 4.5×10<sup>-9</sup> | 0/4 | - | 0.95 | 29.8 (9.6 - 92.8) | 4.5x10<sup>-9</sup> |
| *FGF4* | 0/3 | 2/3 | 61.9 (15.5 - 248.0) | 5.6×10<sup>-9</sup> | 0/0 | - | - | 61.9 (15.5 - 248.0) | 5.6x10<sup>-9</sup> |
| *HINT1* | 1/2 | 1/2 | 66.7 (14.6 - 304.9) | 6.1×10<sup>-8</sup> | 0/1 | - | 0.96 | 66.7 (14.6 - 304.9) | 6.1x10<sup>-8</sup> |
| *PMAIP1* | 0/15 | 3/15 | 22.5 (7.2 - 69.8) | 7.4×10<sup>-8</sup> | 0/4 | - | 0.95 | 22.5 (7.2 - 69.8) | 7.4x10<sup>-8</sup> |
| *PI3* | 0/5 | 2/5 | 42.5 (10.6 - 170.5) | 1.2×10<sup>-7</sup> | 0/0 | - | - | 42.5 (10.6 - 170.5) | 1.2x10<sup>-7</sup> |
| *UBE2V2* | 0/5 | 2/5 | 40.1 (10.0 - 160.9) | 1.9×10<sup>-7</sup> | 0/0 | - | - | 40.1 (10.0 - 160.9) | 1.9x10<sup>-7</sup> |
| *ZNF641* | 2/8 | 0/8 | 49.6 (10.4 - 236.2) | 9.7×10<sup>-7</sup> | 1/1 | 13.9 (0.9 - 224.0) | 6.3×10<sup>-2</sup> | 36.5 (9.4 - 142.5) | 2.2x10<sup>-7</sup> |
| *RHEBL1* | 1/11 | 2/11 | 20.1 (6.4 - 63.8) | 3.2×10<sup>-7</sup> | 0/1 | - | 0.97 | 20.1 (6.4 - 63.8) | 3.2x10<sup>-7</sup> |
| *STK32C* | 0/18 | 3/18 | 18.8 (6.1 - 58.4) | 3.9×10<sup>-7</sup> | 0/1 | - | 0.96 | 18.8 (6.1 - 58.4) | 3.9x10<sup>-7</sup> |
| *INTS15* | 1/3 | 1/3 | 46.3 (10.5 - 203.9) | 4.1×10<sup>-7</sup> | 0/0 | - | - | 46.3 (10.5 - 203.9) | 4.1x10<sup>-7</sup> |
| *TMEM115* | 1/3 | 1/3 | 45.0 (10.2 - 198.4) | 4.9×10<sup>-7</sup> | 0/0 | - | - | 45.0 (10.2 - 198.4) | 4.9x10<sup>-7</sup> |
| *CCL15* | 0/6 | 2/6 | 34.7 (8.7 - 139.1) | 5.4×10<sup>-7</sup> | 0/0 | - | - | 34.7 (8.7 - 139.1) | 5.4x10<sup>-7</sup> |
| *ZNF35* | 1/12 | 2/12 | 17.9 (5.7 - 56.5) | 8.4×10<sup>-7</sup> | 0/0 | - | - | 17.9 (5.7 - 56.5) | 8.4x10<sup>-7</sup> |
| *ACRV1* | 1/26 | 3/2 | 11.9 (4.4 - 32.0) | 8.5×10<sup>-7</sup> | 0/0 | - | - | 11.9 (4.4 - 32.0) | 8.5x10<sup>-7</sup> |
| *PID1* | 0/7 | 2/7 | 32.1 (8.0 - 128.8) | 1.0×10<sup>-6</sup> | 0/0 | - | - | 32.1 (8.0 - 128.8) | 1.0x10<sup>-6</sup> |
| *RHOC* | 1/4 | 1/4 | 37.9 (8.8 - 164.0) | 1.2×10<sup>-6</sup> | 0/2 | - | 0.97 | 37.9 (8.8 - 163.9) | 1.2x10<sup>-6</sup> |
| *PLCG1* | 1/27 | 3/27 | 11.5 (4.3 - 30.9) | 1.2×10<sup>-6</sup> | 0/0 | - | - | 11.5 (4.3 - 30.9) | 1.2x10<sup>-6</sup> |
| *PI15* | 0/7 | 2/7 | 30.0 (7.4 - 121.0) | 1.7×10<sup>-6</sup> | 0/0 | - | - | 30.0 (7.4 - 121.0) | 1.7x10<sup>-6</sup> |
| *PIDD1* | 3/166 | 7/166 | 4.6 (2.5 - 8.6) | 1.6×10<sup>-6</sup> | 1/9 | 1.7 (0.2 - 13.6) | 0.61 | 4.2 (2.3 - 7.7) | 2.1x10<sup>-6</sup> |

When carriers were selected for the presence of both rare PTVs and rare splice-altering variants as predicted by SpliceAI, *MSH6* (RR = 10.9, 95% CI 8.0–14.8), *MLH1* (RR = 25.6, 95% CI 11.4–57.3), *MSH2* (RR = 11.3 95% CI 5.5–23.0 remained the most strongly associated genes, although the relative risk estimates were lower (Supplementary Table 3) compared to the meta-analysis of PTVs only. A total of 18 genes exceeded the exome-wide significance threshold, of which 12 genes were also identified in the PTV only analysis.

### Predicted deleterious missense variants

In the combined meta-analysis of the UKB and 100KG, 9 genes reached exome-wide significance for predicted deleterious MSVs, with odds ratios ranging from 5.7 to 41.3 (Figure 2, Table 2). None of these genes were similarly associated in the PTV only analysis. An additional 35 genes showed associations at p < 0.0001 (Supplementary Table 4). Results from Firth’s regression were similar and the same 9 genes reached exome-wide significance, whereas SKAT-O produced more exaggerated p-values and only 5 of the 9 genes reached exome-wide significance.

**Figure 2.**
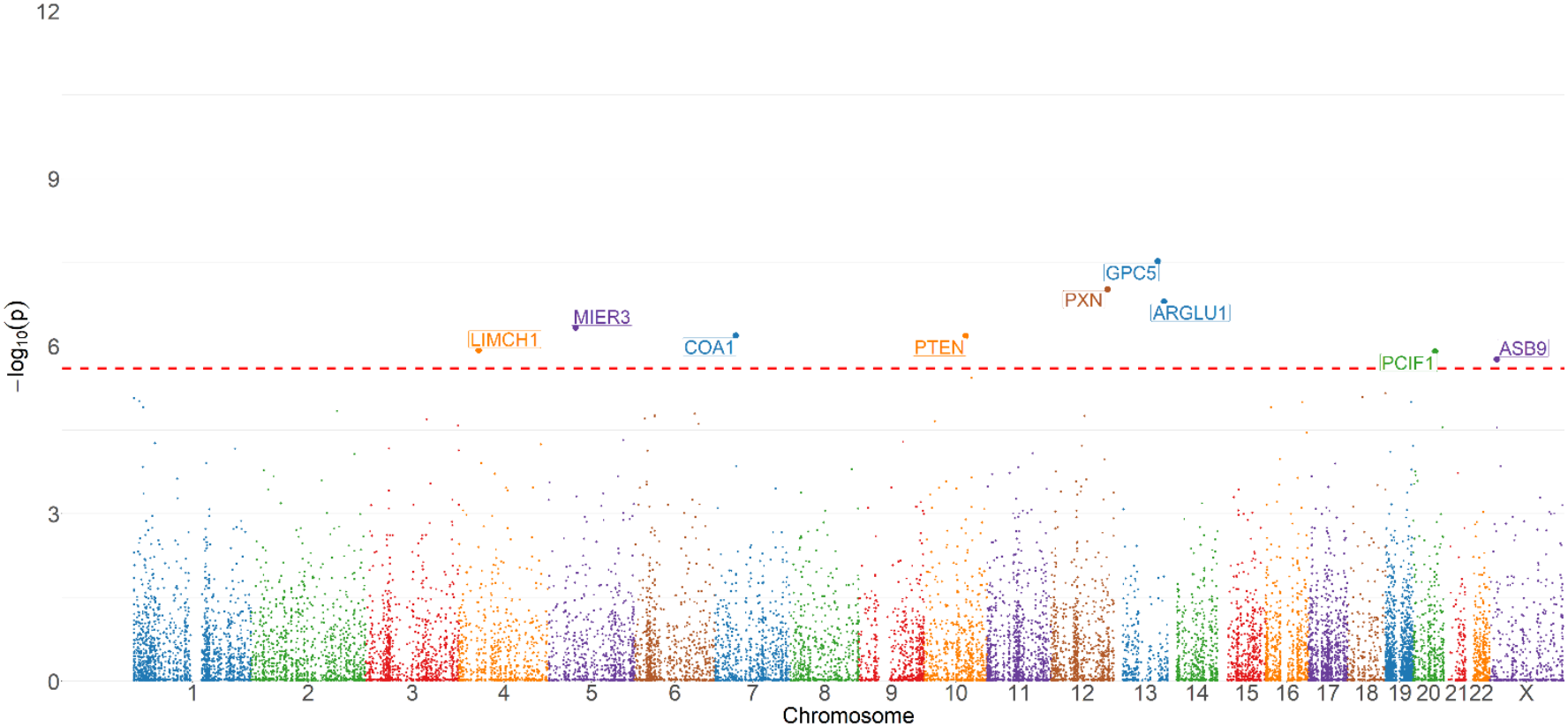
Manhattan plot showing the associations between endometrial cancer and rare predicted deleterious missense variants (MSVs) by gene, based on a meta-analysis combining whole-exome sequencing analysis from the UK Biobank with whole genome sequencing data from 100,000 Genomes Project using logistic regression. The red dashed line represents the exome-wide significance threshold of p=2.5×10−6.

**Table 2.**
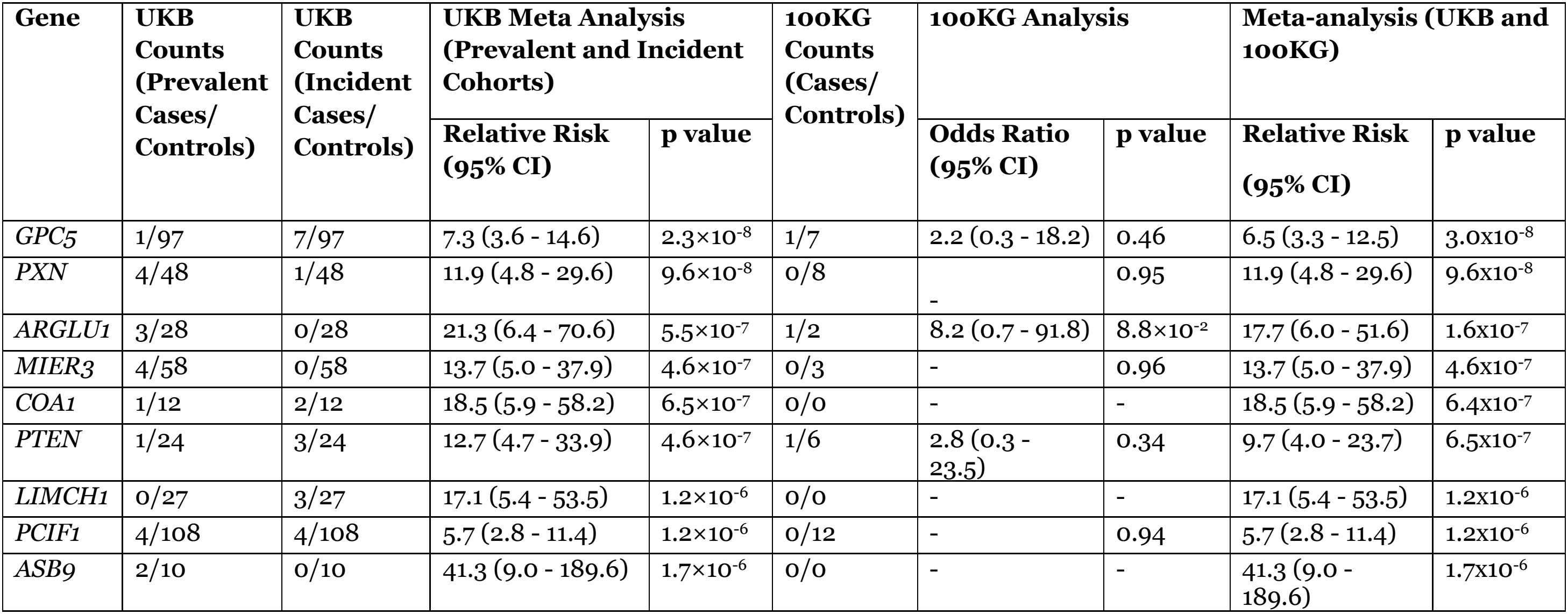
Results of the analysis of rare predicted deleterious missense variants (MSVs) in genes associated with endometrial cancer at exome-wide significance (p < 2.5 × 10^−6^) in the meta-analysis of UK Biobank (UKB) and 100,000 Genomes (100KG) Project datasets using logistic regression. The table shows predicted deleterious MSV carrier counts among cases and controls, together with relative risks (RRs) and 95% confidence intervals (CIs) from the UKB prevalent and incident meta-analyses, the 100KG analysis, and the meta-analyses combining UKB and 100KG.

| Gene | UKB Counts (Prevalent Cases/Controls) | UKB Counts (Incident Cases/Controls) | UKB Meta Analysis (Prevalent and Incident Cohorts) |  | 100KG Counts (Cases/Controls) | 100KG Analysis |  | Meta-analysis (UKB and 100KG) |  |
|---|---|---|---|---|---|---|---|---|---|
|  |  |  | Relative Risk (95% CI) | p value |  | Odds Ratio (95% CI) | p value | Relative Risk (95% CI) | p value |
| *GPC5* | 1/97 | 7/97 | 7.3 (3.6 - 14.6) | 2.3×10<sup>-8</sup> | 1/7 | 2.2 (0.3 - 18.2) | 0.46 | 6.5 (3.3 - 12.5) | 3.0x10<sup>-8</sup> |
| *PXN* | 4/48 | 1/48 | 11.9 (4.8 - 29.6) | 9.6×10<sup>-8</sup> | 0/8 | - | 0.95 | 11.9 (4.8 - 29.6) | 9.6x10<sup>-8</sup> |
| *ARGLU1* | 3/28 | 0/28 | 21.3 (6.4 - 70.6) | 5.5×10<sup>-7</sup> | 1/2 | 8.2 (0.7 - 91.8) | 8.8×10<sup>-2</sup> | 17.7 (6.0 - 51.6) | 1.6x10<sup>-7</sup> |
| *MIER3* | 4/58 | 0/58 | 13.7 (5.0 - 37.9) | 4.6×10<sup>-7</sup> | 0/3 | - | 0.96 | 13.7 (5.0 - 37.9) | 4.6x10<sup>-7</sup> |
| *COA1* | 1/12 | 2/12 | 18.5 (5.9 - 58.2) | 6.5×10<sup>-7</sup> | 0/0 | - | - | 18.5 (5.9 - 58.2) | 6.4x10<sup>-7</sup> |
| *PTEN* | 1/24 | 3/24 | 12.7 (4.7 - 33.9) | 4.6×10<sup>-7</sup> | 1/6 | 2.8 (0.3 - 23.5) | 0.34 | 9.7 (4.0 - 23.7) | 6.5x10<sup>-7</sup> |
| *LIMCH1* | 0/27 | 3/27 | 17.1 (5.4 - 53.5) | 1.2×10<sup>-6</sup> | 0/0 | - | - | 17.1 (5.4 - 53.5) | 1.2x10<sup>-6</sup> |
| *PCIF1* | 4/108 | 4/108 | 5.7 (2.8 - 11.4) | 1.2×10<sup>-6</sup> | 0/12 | - | 0.94 | 5.7 (2.8 - 11.4) | 1.2x10<sup>-6</sup> |
| *ASB9* | 2/10 | 0/10 | 41.3 (9.0 - 189.6) | 1.7×10<sup>-6</sup> | 0/0 | - | - | 41.3 (9.0 - 189.6) | 1.7x10<sup>-6</sup> |

### Absolute risk estimates for well-established genes

The age-specific absolute risks for carriers of PTVs in *MSH6, MLH1*, and *MSH2*, as well as carriers of predicted deleterious MSVs in *PTEN*, are presented in Figure 3. By age 80 years, the estimated absolute risks were 58% (95%CI: 46%–71%), 71% (95% CI: 40%–95%), 27% (95% CI: 13%–49%), and 21% (95% CI: 9%–43%) for *MSH6, MLH1, MSH2* and *PTEN* respectively.

**Figure 3.**
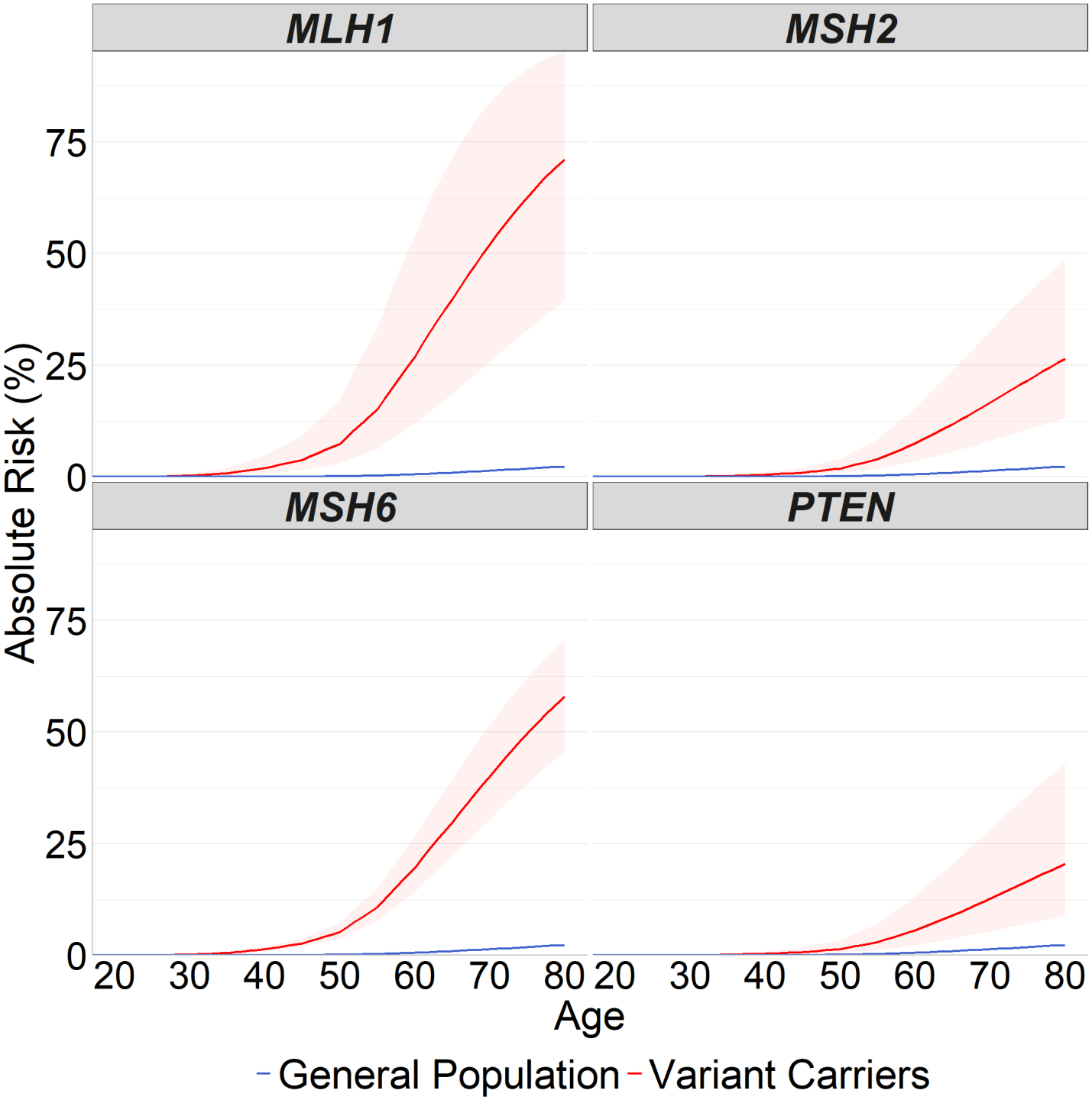
The age-specific absolute risks estimated for carriers of protein truncating variants in *MSH6, MLH1*, and *MSH2*, as well as carriers of deleterious missense variants in *PTEN*. The red lines represent the absolute risk by age for carriers of variants in these genes with 95% confidence interval shown. The blue lines represent the population-level risk for women in the UK population, calculated using 2017-2019 incidence rates from Cancer Research UK.

## DISCUSSION

Here, we conducted a meta-analysis combining whole exome sequencing (WES) data from the UKB with whole genome sequencing (WGS) data from the 100KG, identifying a set of candidate genes associated with endometrial cancer susceptibility.

When considering PTVs only, three well-established susceptibility genes, *MSH6, MLH1* and *MSH2* showed the strongest associations with endometrial cancer. The age-specific absolute endometrial cancer risks for carriers of these PTVs were also broadly consistent with those reported in previous studies ^14,15^. When the selected variants in these genes were filtered even further to include only those annotated as pathogenic or likely pathogenic in the ClinVar database, the results remained similar with slightly larger relative risk estimates (Table 3). For most genes, we did not observe a sufficient number of variant carriers among cases in our dataset to obtain reliable risk estimates. Supplementary Table 5 presents the results for genes with previously reported evidence of association with EC. Notably, the well-established endometrial cancer susceptibility gene *PMS2*^6,16^ did not show exome-wide significant evidence of association with endometrial cancer in our meta-analysis (meta-analysis p = 0.017; RR = 3.72, 95% CI 1.27–10.93) given the low number of PTV carriers in either dataset (Supplementary Table 5). This result remained consistent even after removing variants within the pseudogene overlap region of *PMS2* (meta-analysis p = 0.017; RR = 4.76, 95% CI 1.31–17.22).

**Table 3.** Results for selected genes previously associated with endometrial cancer, analysed using logistic regression. Carrier counts among cases and controls, relative risks (RRs) and 95% confidence intervals (CIs) are presented for rare protein-truncating variants (PTVs) and rare predicted deleterious missense variants (MSVs) classified as pathogenic or likely pathogenic in ClinVar. Results are shown separately for the UKB prevalent and incident meta-analyses, the 100KG analysis, and the meta-analyses combining UKB and 100KG.

| **Gene** | **UKB Counts (Prevalent Cases/Controls)** | **UKB Counts (Incident Cases/Controls)** | **UKB Meta Analysis (Prevalent and Incident Cohorts)** |  | **100K Genomes Counts (Cases/Controls)** | **100KG Analysis** |  | **Meta-analysis (UKB and 100KG)** |  |
|---|---|---|---|---|---|---|---|---|---|
|  |  |  | **Relative Risk (95% CI)** | **p value** |  | **Odds Ratio (95% CI)** | **p value** | **Relative Risk (95% CI)** | **p value** |
| **Protein-truncating variants (PTVs)** |  |  |  |  |  |  |  |  |  |
| *MSH6* | 16/60 | 16/60 | 38.3 (26.5 - 55.5) | 2.3×10<sup>-83</sup> | 7/2 | 63.4 (12.9 - 311.0) | 3.2×10<sup>-7</sup> | 39.3 (27.5 - 56.4) | 5.7x10<sup>-89</sup> |
| *MLH1* | 4/7 | 1/7 | 65.0 (22.8 - 185.4) | 5.6×10<sup>-15</sup> | 0/1 | - | 0.96 | 65.0 (22.8 - 185.3) | 5.6x10<sup>-15</sup> |
| *MSH2* | 4/0 | 1/0 | 257.1 (36.0 - 1837.4) | 3.2×10<sup>-8</sup> | 1/3 | 5.1 (0.5 - 49.6) | 0.16 | 50.0 (10.9 - 212.0) | 3.3x10<sup>-7</sup> |
| **Missense variants (MSVs) predicted to be deleterious** |  |  |  |  |  |  |  |  |  |
| *MSH6* | 2/11 | 3/11 | 32.0 (12.9 - 79.2) | 7.1×10<sup>-14</sup> | 2/1 | 31.9 (2.8 - 358.6) | 5.0×10<sup>-3</sup> | 32.0 (13.7 - 74.8) | 1.3x10<sup>-15</sup> |
| *MLH1* | 3/2 | 0/2 | 303.5 (50.0 - 1842.0) | 5.2×10<sup>-10</sup> | 3/2 | 26.5 (4.3 - 162.0) | 4.0×10<sup>-3</sup> | 90.1 (25.1 - 323.6) | 5.1x10<sup>-12</sup> |
| *PTEN* | 0/6 | 1/6 | 18.9 (2.7 - 134.2) | 3.3×10<sup>-3</sup> | 0/1 | - | 0.96 | 18.9 (2.7 - 134.1) | 3.3x10<sup>-3</sup> |
| *MSH2* | 0/5 | 0/5 | - | 0.96 | 0/0 | - | - | - | - |

In addition to *MSH6, MLH1* and *MSH2*, we identified 19 additional genes reaching exome-wide significance (p < 2.5 × 10^−6^) for association with endometrial cancer (Figure 1, Table 1). We identified a further six genes when PTVs were analysed together with splice-altering variants as predicted by SpliceAI (Supplementary Table 3). Among the genes with evidence at exome-wide significance, *PHKG2, SUPT16H* and *FNDC1* have previously been implicated in endometrial cancer ^17–19^, while *HINT1, PMAIP1, ERMN, STK32C, ACRV1, RHOC, PLCG1*, and *PIDD1* have been implicated in colorectal cancer ^20–27^. Some of the additional genes we identified have also been implicated in other cancers or tumour-related processes, though evidence remains preliminary (Supplementary Table 2). For example, *FGF4* encodes fibroblast growth factor 4 which has been linked to increased proliferation and metastasis in breast cancer cells ^28^, and *PID1* is a tumour suppressor gene found to be associated with apoptosis and inhibition of tumour growth in embryonal brain tumours, gliomas and bladder cancer ^29,30^. A further 48 genes showed suggestive associations with endometrial cancer risk at p < 0.0001, of which three genes have had some prior evidence suggestive of an association with endometrial cancer, nine with colorectal cancer, and 25 with other cancers (Supplementary Table 2).

When considering predicted deleterious MSVs, we confirmed the well-known association with *PTEN* ^31–33^. The association was not identified in the PTV analysis, where no *PTEN* PTV carriers were observed among UK Biobank cases, although two were identified among cases in the 100K Genomes dataset. The number of carriers was too small to compare EC risk between the two variant classes. The age-specific absolute endometrial cancer risks for carriers of these MSVs in *PTEN* were also consistent with those reported in previous studies of individuals with *PTEN* Hamartoma Tumor Syndrome (PHTS) ^34,35^. However, this association was no longer observed after restricting the analysis to *PTEN* variants classified as pathogenic or likely pathogenic in the ClinVar database (Table 3), likely because the ClinVar filtering substantially reduced the number of eligible variants (Supplementary Table 6), leaving only one endometrial cancer carrier in the dataset. We identified eight additional genes associated with endometrial cancer risk at exome-wide significance (Figure 2, Table 2). Although these genes have previously been implicated in various cancers all with previously reported but unconfirmed associations to various cancers, their associations with endometrial cancer risk have not been confirmed (Supplementary Table 4). Notably, the genes *PXN, COA1, ASB9* and *PCIF1* have some suggestive evidence of associations to colorectal cancer, while the remaining four genes have been implicated in a risk of other cancers (Supplementary Table 4).

The novel candidate predisposition genes identified in our analysis showed high-risk associations with relative risks ranging from 4.2 to 66.7 for PTVs (Table 1) and 5.7 to 41.3 for MSVs (Table 2). Given the rarity of the variants included in this analysis and the small sample size, the risk estimates for some genes remain imprecise, as reflected by their wide confidence intervals. The lower confidence limits provide a more conservative indication of the potential magnitude of risk, and the estimates should be interpreted with caution until replicated in independent studies. To account for potential bias arising from the small number of carriers, we applied Firth’s bias-reduced regression, which yielded results highly consistent with those from standard logistic regression (Supplementary Tables 2 and 4). Simple burden tests are most powerful when all variants have similar effect size, an assumption that is plausible for PTVs which typically lead to loss of function but may be less appropriate for MSVs. Therefore, we additionally performed SKAT-O which should be more powerful when there is heterogeneity in variant effect sizes or directions. As SKAT-O requires at least two carriers of the same variant, only a subset of genes met this criterion (Supplementary Table 4).

When PTVs were analysed together with splice-altering variants predicted by SpliceAI, the overall results remained consistent, although the estimated odds ratios were attenuated. While PTVs generally lead to clear loss of function and higher penetrance, SpliceAI-predicted splice variants include both true splice-disrupting events and variants with partial or minimal effects on splicing. The addition of such variants increases the number of carriers, particularly within the control population, thereby diluting the strong association signal observed for PTVs alone.

This study has several limitations. First, although it combines two of the largest publicly available germline sequencing cohorts in the UK, the number of endometrial cancer cases, particularly within the 100KG cohort, remains modest. This limits the statistical power necessary to detect very rare variant associations. Consequently, the observed associations and risk estimates should be interpreted with caution, as confidence intervals remain wide. Second, differences in variant calling pipelines and quality control thresholds between the UKB and 100KG may also have hindered the harmonization of results across the two datasets, even though the per-gene heterogeneity between the two studies was not significant (Supplementary Tables 2 and 4). Third, analyses were restricted to women of European ancestry because of the limited number of cases in other ethnic groups (n = 128 in the UKB; n = 86 in the 100KG), which may reduce the generalizability of these findings. Therefore, replication in larger and more diverse cohorts will be required to confirm these associations and refine effect size estimates.

In summary, this study expands the landscape of genetic susceptibility to endometrial cancer by highlighting novel candidate genes that warrant replication in larger cohorts. However, these associations require rigorous confirmation to obtain reliable, unbiased risk estimates before they can inform clinical translation.

## METHODS

### Datasets

#### UK Biobank

UKB recruited over 500,000 individuals across the UK between 2005 and 2010. WES for UKB participants was generated in collaboration with biopharmaceutical companies, and the final data was released in July 2022 for access via the UKB Research Analysis Platform. Quality control and processing of the WES data is described in detail in Szustakowski et al. 2021^36^. Our analysis began with the Variant Call Format (VCF) files generated from this protocol. Our inclusion criteria entailed selecting for self-identified women with available WES data and no reported history of tamoxifen use. For the control group, we also selected for those who had not undergone a hysterectomy. To restrict the analysis to unrelated individuals, we excluded women with more than ten relatives and retained only one participant from each group of first-degree relatives (Supplementary Figure 1). We restricted the analysis to women of European ancestry, whose self-reported ethnicity was in accordance with genetically inferred ancestry using iAdmix ^37^. Endometrial cancer cases were identified based on a recorded primary diagnosis with the ICD-10 code C54 and were analysed separately as prevalent or incident cases. Incident cases were women diagnosed with endometrial cancer more than one year after recruitment. Of the 148,948 controls initially identified in UKB, 31 were excluded because they were recorded as deceased within one year of recruitment, leaving 148,917 controls for the Cox regression analysis. All UKB participants provided informed consent. The UKB has approval from the North West Multi-centre Research Ethics Committee as a Research Tissue Bank approval.

#### 100,000 Genomes

The 100KG Project recruited over 85,000 NHS patients affected by rare diseases or cancer with WGS completed in 2018. The genome delivery protocols used to process the WGS data is described in the accompanying information for the latest release (version 19, October 2024). Our analysis began with VCF files generated from the relevant Genomics England and Illumina genome delivery protocols for this release. We selected cases as women diagnosed with only endometrial cancer from the Cancer Programme, and controls as unaffected relatives of participants in the Rare Disease Programme. Eligible participants were restricted to females with karyotype XX and European ancestry, with one control per family retained in the following order of priority: siblings, aunts, grandmothers, then mothers (Supplementary Figure 2). Research ethics governance is in place for the 100,000 Genomes Project via independent Health Research Authority ethics approval and scrutiny, as well as Genomic England’s corporate governance, including the Ethics Advisory Committee which advises the Board of Directors of Genomics England.

### Variant Selection and Annotation

We applied consistent quality control filters to VCF files from both the UKB and 100KG Project. Single Nucleotide Polymorphisms (SNPs) with sequencing depth < 7 and insertions or deletions (InDels) with depth < 10 were excluded. Rare variants were selected as those with a minor allele frequency <0.001 calculated using PLINK v1.90(Purcell et al., 2007) in both the control groups, and across all seven gnomAD v4.0 populations (African/African-American, Ashkenazi Jewish, East Asian, South Asian, Finnish, Non-Finnish European, and Middle Eastern).

The selected variants were annotated using the Ensembl Variant Effect Predictor (VEP) v112 ^38^ with the AlphaMissense ^39^ and SpliceAI ^40^ plug-ins. The MANE Select transcript was used when available, otherwise the RefSeq Select transcript was substituted. PTVs were defined as frameshift, stop loss, stop gained, start loss, and splice region variants using VEP annotation. Those located in the last exon or within the last 50 bp of the penultimate exon were excluded, as these variants are expected to escape nonsense-mediated mRNA decay. As a sensitivity analysis, we also included any splice-altering variants with a SpliceAI delta score >0.5 (acceptor loss, acceptor gain, donor loss or donor gain). Predicted deleterious MSVs were defined as those annotated as “likely pathogenic” by AlphaMissense. In an additional analysis, both protein truncating and missense variants in genes already known to be associated with endometrial cancer susceptibility, including *MSH2, MSH6, MLH1, PMS2* and *PTEN*, were filtered to keep only those annotated as pathogenic and/or likely pathogenic in the ClinVar database (Supplementary Table 6). In this analysis, variants in the exons 9, 11, 12, 13, 14 and 15 of the gene *PMS2* were also excluded to mitigate interference from overlapping pseudogenes.

### Gene Burden Analysis

To test associations between rare variants and endometrial cancer risk, we performed gene-based burden tests in which variants within each gene were collapsed. Analyses were restricted to genes with at least four carriers of any variant selected to be of interest. Participant genotypes were coded as 0/1 to indicate the absence or presence of a qualifying variant within a gene. All models were adjusted for the first ten ancestry informative principal components provided by the UKB. For the UKB prospective cohort, associations were evaluated using Cox proportional hazards models to account for time-to-event to estimate a hazard ratio associated with carrier status. For the UKB retrospective cohort and the 100KG dataset, logistic regression was used to estimate the odds ratio associated with carrier status. Because an incident diagnosis of endometrial cancer was rare in the UK Biobank cohort (incidence rate 0.57 per 1,000 person-years; cumulative incidence 0.8% over the 14 years of follow-up), odds ratios from logistic regression were assumed to closely approximate hazard ratios from the time-to-event analyses. Meta-analyses were then conducted to first combine results from the UKB prevalent and incident datasets based on the assumption that the hazard ratio and odds ratio estimate the same underlying relative risk (RR), followed by a second meta-analysis combining results from the UKB and the 100KG Project. To explore alternative approaches to standard logistic regression, we additionally applied Firth’s bias-reduced logistic regression to account for potential small-sample bias ^41^ in each dataset separately, before meta-analysis, in the 100KG and the prevalent UKB cohort. We also conducted gene-based association testing using SKAT-O, which allows for variation in effect sizes or directions among variants within a gene ^42^ for predicted deleterious MSVs.

Absolute risks for carriers were estimated by scaling UK age-specific endometrial cancer incidence rates for 2017–2019 (Cancer Research UK Stats Data Hub)^43^ by estimated relative risks (RRs) and confidence bounds and converting to cumulative risk using 1 − exp(−∑λ_t_).

## Data Availability

Requests for UK Biobank should be made to the UK Biobank Access Management Team.

## Acknowledgements

This work is supported by Cancer Research UK Early Detection and Diagnosis Award: EDDPMA-May23/100008. X.Y. and A.C.A. are supported by Cancer Research UK grant: PPRPGM-Nov20\100002. A.C.A. is supported by Cancer Research UK grant: SEBCD3-2024/100001. E.J.D. is supported by a National Institute for Health and Care Research (NIHR) Research Professorship (NIHR304303) and the NIHR Manchester Biomedical Research Centre (NIHR203308). M.T. was supported by the NIHR Cambridge Biomedical Research Centre (NIHR203312). Tracy A. O’Mara is supported by a National Health and Medical Research Council (NHMRC) of Australia Investigator Fellowship (#ID 2041819).

## Author contributions

Conceptualization: X.Y., A.C.A., D.F.E.; Methodology: X.Y., A.C.A, D.F.E, T.A.O., A.B.S; Formal analysis: M.L., X.Y.; Investigation: X.Y., A.C.A, D.F.E, T.A.O., A.B.S, M.T., P.D.P.P, E.J.D, M.L.; Data curation: M.L.; Writing-original draft preparation: M.L., X.Y.; Writing-review and editing: All authors reviewed and edited the paper. All authors are responsible for the decision to submit for publication. All authors agreed to the published version of the manuscript.

