## Supplementary Material for "Meta-analysis of Sequencing Data Identifies Candidate Endometrial Cancer Susceptibility Genes"

### SUPPLEMENTARY INFO

Supplementary Table 1. Demographic Characteristics of the UK Biobank (UKB) and 100K Genomes (100KG) Study Groups.

| | UK Biobank | | | 100K Genomes | |
|---|---|---|---|---|---|
| | Prevalent Cases, No. % | Incident Cases, No. % | Controls, No. % | Cases, No. % | Controls, No. % |
| Age groups (years) | | | | | |
| 0 - 9 | 0 (0.0) | 0 (0.0) | 0 (0.0) | 0 (0.0) | 32 (0.27) |
| 10 - 19 | 0 (0.0) | 0 (0.0) | 0 (0.0) | 0 (0.0) | 43 (0.47) |
| 20 - 29 | 0 (0.0) | 0 (0.0) | 0 (0.0) | 1 (0.14) | 1,254 (10.71) |
| 30 - 39 | 14 (1.86) | 0 (0.0) | 0 (0.0) | 9 (1.25) | 4,139 (35.36) |
| 40 - 49 | 94 (12.5) | 23 (1.91) | 62 (0.04) | 24 (3.34) | 3,350 (28.62) |
| 50 - 59 | 389 (51.73) | 225 (18.69) | 22,883 (15.36) | 137 (19.05) | 1,539 (13.15) |
| 60 - 69 | 254 (33.78) | 556 (46.18) | 50,000 (33.57) | 226 (31.43) | 753 (6.43) |
| 70 - 79 | 1 (0.13) | 388 (32.23) | 60,892 (40.88) | 241 (33.52) | 457 (3.90) |
| 80 - 89 | 0 (0.0) | 12 (0.99) | 15,111 (10.15) | 75 (10.43) | 127 (1.08) |
| 90 - 99 | 0 (0.0) | 0 (0.0) | 0 (0.0) | 6 (0.83) | 12 (0.10) |
| Total | 752 | 1,204 | 148,948 | 719 | 11,706 |
| Mean (sd) | 56.7 (± 6.5) | 66 (± 7.2) | 69.1 (± 8.0) | 67.4 (± 10.6) | 42.9 (± 13.0) |
| Median (IQR) | 57.2 (53.4 - 61.1) | 66.7 (61.3 - 71.4) | 70 (63 - 76) | 68 (60 - 75) | 40 (34 - 49) |

Supplementary Table 2. Genes with rare protein-truncating variants (PTVs) associated with $p < 10^{-4}$ in the meta-analysis combining whole-exome sequencing data from the UK Biobank (UKB) and whole-genome sequencing data from the 100,000 Genomes Project (100KG). Results are presented for meta-analyses using standard logistic regression and Firth's bias-reduced regression. Heterogeneity between UKB and 100KG was assessed using Cochran's Q statistic and the $I^2$ statistic. Previous evidence of associations with endometrial, colorectal or other cancers is also reported.

| **Gene** | **Logistic regression meta-analysis** | | **Firth's regression meta-analysis** | | **Heterogeneity** | | | **Previous cancer evidence** |
|---|---|---|---|---|---|---|---|---|
| | **p value** | **Relative Risk (95% CI)** | **p value** | **Relative Risk (95% CI)** | **Q statistic** | **p value of Q statistic** | **$I^2$** | |
| *MSH6* | $2.2 \times 10^{-92}$ | 36.6 (25.9 - 51.8) | $2.9 \times 10^{-93}$ | 36.8 (26.1 - 52.0) | $4.3 \times 10^{-1}$ | 0.5 | 0 | Endometrial[1] |
| *MLH1* | $8.1 \times 10^{-18}$ | 52.5 (21.3 - 129.4) | $2.1 \times 10^{-18}$ | 54.8 (22.4 - 134.3) | $4.3 \times 10^{-3}$ | 0.9 | 0 | Endometrial[1,2] |
| *MSH2* | $7.6 \times 10^{-11}$ | 13.0 (6.0 - 28.2) | $3.9 \times 10^{-12}$ | 14.4 (6.8 - 30.5) | $1.7 \times 10^{-1}$ | 0.7 | 0 | Endometrial[1,2] |
| *PHKG2* | $4.5 \times 10^{-9}$ | 29.8 (9.6 - 92.8) | $4.5 \times 10^{-9}$ | 29.8 (9.6 - 92.8) | $6.6 \times 10^{-3}$ | 0.9 | 0 | Endometrial[3,4], Other[5] |
| *FGF4* | $5.6 \times 10^{-9}$ | 61.9 (15.5 - 248.0) | $5.6 \times 10^{-9}$ | 61.9 (15.5 - 248.0) | 0 | - | - | Other[6–9] |
| *HINT1* | $6.1 \times 10^{-8}$ | 66.7 (14.6 - 304.9) | $6.5 \times 10^{-5}$ | 118.4 (11.4 - 1231.9) | $4.4 \times 10^{-3}$ | 0.9 | 0 | Colorectal[10,11] |
| *PMAIP1* | $7.4 \times 10^{-8}$ | 22.5 (7.2 - 69.8) | $7.4 \times 10^{-8}$ | 22.5 (7.2 - 69.8) | $6.3 \times 10^{-3}$ | 0.9 | 0 | Colorectal[12,13] |
| *PI3* | $1.3 \times 10^{-7}$ | 42.5 (10.6 - 170.5) | $1.3 \times 10^{-7}$ | 42.5 (10.6 - 170.6) | 0 | - | - | - |
| *UBE2V2* | $1.9 \times 10^{-7}$ | 40.1 (10.0 - 160.9) | $1.9 \times 10^{-7}$ | 40.1 (10.0 - 160.9) | 0 | - | - | Other[14,15] |
| *ZNF641* | $2.2 \times 10^{-7}$ | 36.5 (9.4 - 142.5) | $2.4 \times 10^{-8}$ | 42.0 (11.3 - 156.2) | $6.1 \times 10^{-1}$ | 0.4 | 0 | Other[16,17] |
| *RHEBL1* | $3.3 \times 10^{-7}$ | 20.1 (6.4 - 63.8) | $2.3 \times 10^{-8}$ | 22.8 (7.6 - 68.1) | $3.3 \times 10^{-3}$ | 1.0 | 0 | Other[18,19] |
| *STK32C* | $3.9 \times 10^{-7}$ | 18.8 (6.1 - 58.4) | $3.9 \times 10^{-7}$ | 18.8 (6.1 - 58.4) | $3.5 \times 10^{-3}$ | 1.0 | 0 | Colorectal[20] |
| *INTS15* | $4.1 \times 10^{-7}$ | 46.3 (10.5 - 203.9) | $8.0 \times 10^{-8}$ | 52.5 (12.4 - 223.2) | 0 | - | - | - |
| *TMEM115* | $4.9 \times 10^{-7}$ | 45.0 (10.2 - 198.4) | $9.8 \times 10^{-8}$ | 51.1 (12.0 - 217.2) | 0 | - | - | Other[21–23] |
| *CCL15* | $5.4 \times 10^{-7}$ | 34.7 (8.7 - 139.1) | $5.4 \times 10^{-7}$ | 34.7 (8.7 - 139.1) | 0 | - | - | Other[24–26] |
| *ZNF35* | $8.4 \times 10^{-7}$ | 17.9 (5.7 - 56.5) | $6.1 \times 10^{-8}$ | 20.3 (6.8 - 60.4) | 0 | - | - | Other[27,28] |
| *ACRV1* | $8.5 \times 10^{-7}$ | 11.9 (4.4 - 32.0) | $9.1 \times 10^{-8}$ | 13.0 (5.1 - 33.2) | 0 | - | - | Colorectal[29,30] |
| *PID1* | $1.0 \times 10^{-6}$ | 32.1 (8.0 - 128.8) | $1.0 \times 10^{-6}$ | 32.1 (8.0 - 128.8) | 0 | - | - | Other[31,32] |
| *RHOC* | $1.2 \times 10^{-6}$ | 37.9 (8.8 - 163.9) | $1.4 \times 10^{-7}$ | 44.3 (10.8 - 181.8) | $3.5 \times 10^{-3}$ | 1.0 | 0 | Colorectal[33,34] |

| | | | | | | | | |
|---|---|---|---|---|---|---|---|---|
| *PLCG1* | $1.2x10^{-6}$ | 11.5 (4.3 - 30.9) | $1.6x10^{-7}$ | 12.3 (4.8 - 31.4) | 0 | - | - | Colorectal[35,36], Other[37,38] |
| *PI15* | $1.7x10^{-6}$ | 30.0 (7.4 - 121.0) | $1.7x10^{-6}$ | 30.0 (7.4 - 121.0) | 0 | - | - | - |
| *PIDD1* | $2.1x10^{-6}$ | 4.2 (2.3 - 7.7) | $3.9x10^{-7}$ | 4.5 (2.5 - 7.9) | $8.0x10^{-1}$ | 0.4 | 0 | Colorectal[39], Other[40,41] |
| *CLIP2* | $2.8x10^{-6}$ | 15.0 (4.8 - 46.6) | $2.8x10^{-6}$ | 15.0 (4.8 - 46.6) | $3.4x10^{-3}$ | 1.0 | 0 | Other[42] |
| *PRPF18* | $2.9x10^{-6}$ | 37.1 (8.2 - 168.4) | $1.5x10^{-7}$ | 44.6 (10.8 - 183.8) | 0 | - | - | - |
| *PYCR2* | $2.9x10^{-6}$ | 27.5 (6.9 - 110.1) | $2.9x10^{-6}$ | 27.5 (6.9 - 110.1) | 0 | - | - | Colorectal[43,44] |
| *ZNF100* | $2.9x10^{-6}$ | 15.0 (4.8 - 46.5) | $2.9x10^{-6}$ | 15.0 (4.8 - 46.5) | $2.9x10^{-3}$ | 1.0 | 0 | Other[45,46] |
| *MEST* | $3.1x10^{-6}$ | 27.2 (6.8 - 108.8) | $3.1x10^{-6}$ | 27.2 (6.8 - 108.8) | 0 | - | - | Other[47–49] |
| *PIP5K1A* | $3.2x10^{-6}$ | 11.1 (4.0 - 30.7) | $2.5x10^{-7}$ | 12.8 (4.9 - 33.8) | $3.2x10^{-3}$ | 1.0 | 0 | Colorectal[50,51] |
| *CLCF1* | $4.9x10^{-6}$ | 29.3 (6.9 - 124.9) | $4.9x10^{-7}$ | 34.8 (8.7 - 139.0) | $3.9x10^{-3}$ | 1.0 | 0 | Other[52,53] |
| *KCNA7* | $5.8x10^{-6}$ | 15.2 (4.7 - 49.3) | $4.3x10^{-7}$ | 17.9 (5.9 - 55.0) | $3.0x10^{-3}$ | 1.0 | 0 | Other[54] |
| *AGTPBP1* | $5.8x10^{-6}$ | 13.8 (4.4 - 42.8) | $5.8x10^{-6}$ | 13.8 (4.4 - 42.8) | $2.9x10^{-3}$ | 1.0 | 0 | Other[55,56] |
| *SEC11A* | $6.4x10^{-6}$ | 24.4 (6.1 - 97.7) | $6.4x10^{-6}$ | 24.4 (6.1 - 97.7) | 0 | - | - | Other[57,58] |
| *MBD1* | $7.0x10^{-6}$ | 24.1 (6.0 - 96.5) | $7.0x10^{-6}$ | 24.1 (6.0 - 96.6) | $3.7x10^{-3}$ | 1.0 | 0 | Other[59–61] |
| *IGSF5* | $7.0x10^{-6}$ | 5.1 (2.5 - 10.4) | $7.3x10^{-7}$ | 5.5 (2.8 - 10.9) | 1.8 | 0.2 | 0.4 | Colorectal[62] |
| *RERG* | $7.8x10^{-6}$ | 29.4 (6.7 - 129.4) | $4.0x10^{-7}$ | 35.5 (8.9 - 141.0) | 0 | - | - | Other[63–65] |
| *ADCY4* | $8.6x10^{-6}$ | 3.5 (2.0 - 6.1) | $2.1x10^{-6}$ | 3.7 (2.2 - 6.3) | $1.8x10^{-2}$ | 0.9 | 0 | Other[66–68] |
| *IGSF3* | $9.3x10^{-6}$ | 7.4 (3.1 - 17.9) | $1.3x10^{-6}$ | 8.2 (3.5 - 19.1) | $2.7x10^{-3}$ | 1.0 | 0 | Other[69–71] |
| *PPFIA2* | $9.5x10^{-6}$ | 23.0 (5.7 - 92.0) | $9.5x10^{-6}$ | 23.0 (5.7 - 92.0) | $3.3x10^{-3}$ | 1.0 | 0 | Other[72,73] |
| *DYRK4* | $1.1x10^{-5}$ | 27.7 (6.3 - 121.9) | $1.4x10^{-6}$ | 10.3 (4.0 - 26.5) | $4.1x10^{-1}$ | 0.5 | 0 | Other[74,75] |
| *ZNF169* | $1.1x10^{-5}$ | 9.4 (3.5 - 25.5) | $6.0x10^{-7}$ | 33.5 (8.4 - 133.0) | $6.4x10^{-3}$ | 0.9 | 0 | Colorectal[76] |
| *LZIC* | $1.2x10^{-5}$ | 27.8 (6.3 - 122.9) | $7.4x10^{-7}$ | 33.3 (8.3 - 133.4) | $3.8x10^{-3}$ | 1.0 | 0 | Other[77] |
| *XKRX* | $1.2x10^{-5}$ | 25.2 (6.0 - 106.8) | $9.6x10^{-7}$ | 30.5 (7.8 - 119.5) | 0 | - | - | - |
| *TMEM38B* | $1.6x10^{-5}$ | 8.7 (3.3 - 23.4) | $2.4x10^{-6}$ | 9.5 (3.7 - 24.2) | $2.8x10^{-3}$ | 1.0 | 0 | Other[78] |
| *TMEM14A* | $1.7x10^{-5}$ | 21.1 (5.3 - 84.7) | $1.7x10^{-5}$ | 21.1 (5.3 - 84.7) | 0 | - | - | Other[79,80] |
| *FANCD2OS* | $1.8x10^{-5}$ | 23.4 (5.5 - 98.7) | $1.6x10^{-6}$ | 28.3 (7.2 - 111.0) | 0 | - | - | - |
| *PIMREG* | $1.8x10^{-5}$ | 8.6 (3.2 - 22.9) | $1.8x10^{-5}$ | 8.6 (3.2 - 22.9) | 0 | - | - | Other[81–83] |
| *RDX* | $2.0x10^{-5}$ | 11.8 (3.8 - 36.7) | $1.9x10^{-5}$ | 11.8 (3.8 - 36.7) | $4.2x10^{-3}$ | 0.9 | 0 | Other[84] |
| *NFKBID* | $2.0x10^{-5}$ | 20.6 (5.1 - 82.4) | $2.0x10^{-5}$ | 20.6 (5.1 - 82.4) | 0 | - | - | Other[85,86] |

| | | | | | | | | |
|---|---|---|---|---|---|---|---|---|
| *NDUFS1* | $2.0 \times 10^{-5}$ | 8.4 (3.2 - 22.6) | $2.0 \times 10^{-5}$ | 8.5 (3.2 - 22.6) | $2.7 \times 10^{-3}$ | 1.0 | 0 | Colorectal[87,88] |
| *TRIM23* | $2.1 \times 10^{-5}$ | 24.1 (5.6 - 104.2) | $9.7 \times 10^{-7}$ | 29.3 (7.6 - 113.1) | 0 | - | - | Colorectal[89–91] |
| *ORC3* | $2.2 \times 10^{-5}$ | 4.0 (2.1 - 7.6) | $6.8 \times 10^{-6}$ | 4.2 (2.2 - 7.7) | $1.7 \times 10^{-4}$ | 1.0 | 0 | Other[92–94] |
| *CCNI* | $2.4 \times 10^{-5}$ | 19.9 (5.0 - 79.7) | $2.4 \times 10^{-5}$ | 19.9 (5.0 - 79.7) | $3.6 \times 10^{-3}$ | 1.0 | 0 | Other[95–97] |
| *SETDB2* | $2.5 \times 10^{-5}$ | 11.4 (3.7 - 35.5) | $2.5 \times 10^{-5}$ | 11.4 (3.7 - 35.5) | $4.3 \times 10^{-3}$ | 0.9 | 0 | Other[98–100] |
| *OFD1* | $3.0 \times 10^{-5}$ | 19.2 (4.8 - 77.0) | $3.0 \times 10^{-5}$ | 19.2 (4.8 - 77.0) | 0 | - | - | Endometrial[101] |
| *WDR19* | $3.7 \times 10^{-5}$ | 3.7 (2.0 - 6.9) | $1.1 \times 10^{-5}$ | 3.9 (2.1 - 7.2) | $4.6 \times 10^{-3}$ | 0.9 | 0 | Other[102,103] |
| *FNDC1* | $3.8 \times 10^{-5}$ | 5.4 (2.4 - 12.1) | $8.4 \times 10^{-6}$ | 5.8 (2.7 - 12.7) | $4.4 \times 10^{-3}$ | 0.9 | 0 | Endometrial[104], Colorectal[105,106], Other[107,108] |
| *LY6L* | $4.1 \times 10^{-5}$ | 12.9 (3.8 - 44.0) | $1.6 \times 10^{-6}$ | 16.1 (5.2 - 50.1) | $4.5 \times 10^{-1}$ | 0.5 | 0 | Endometrial[109], Other[110] |
| *MYPN* | $4.4 \times 10^{-5}$ | 7.9 (2.9 - 21.4) | $5.1 \times 10^{-6}$ | 9.1 (3.5 - 23.4) | $2.6 \times 10^{-3}$ | 1.0 | 0 | - |
| *TP53BP1* | $5.0 \times 10^{-5}$ | 11.2 (3.5 - 35.9) | $4.5 \times 10^{-6}$ | 12.9 (4.3 - 38.6) | $2.8 \times 10^{-3}$ | 1.0 | 0 | Colorectal[111] |
| *TOP6BL* | $5.6 \times 10^{-5}$ | 12.3 (3.6 - 41.8) | $6.7 \times 10^{-6}$ | 13.2 (4.3 - 40.7) | 1.2 | 0.3 | 0.2 | - |
| *SLC11A1* | $5.8 \times 10^{-5}$ | 6.1 (2.5 - 14.6) | $1.7 \times 10^{-5}$ | 6.3 (2.7 - 14.6) | $2.8 \times 10^{-3}$ | 1.0 | 0 | Colorectal[112–114] |
| *EXOSC10* | $5.9 \times 10^{-5}$ | 5.5 (2.4 - 12.5) | $5.4 \times 10^{-6}$ | 6.3 (2.8 - 13.8) | $7.1 \times 10^{-5}$ | 1.0 | 0 | Colorectal[115,116] |
| *DHRS4* | $6.1 \times 10^{-5}$ | 7.6 (2.8 - 20.6) | $6.8 \times 10^{-6}$ | 8.8 (3.4 - 22.6) | $5.3 \times 10^{-3}$ | 0.9 | 0 | Other[117–119] |
| *SENP7* | $6.2 \times 10^{-5}$ | 8.1 (2.9 - 22.7) | $6.9 \times 10^{-5}$ | 7.9 (2.9 - 22.0) | 1.7 | 0.2 | 0.4 | Colorectal[120,121] |
| *PLA2G2A* | $6.3 \times 10^{-5}$ | 10.8 (3.4 - 34.9) | $6.0 \times 10^{-6}$ | 12.5 (4.2 - 37.4) | $2.9 \times 10^{-3}$ | 1.0 | 0 | Colorectal[122] |
| *ECM2* | $7.1 \times 10^{-5}$ | 16.8 (4.2 - 67.7) | $7.4 \times 10^{-5}$ | 16.7 (4.1 - 67.1) | 0 | - | - | Other[123–125] |
| *CTDP1* | $7.8 \times 10^{-5}$ | 7.7 (2.8 - 21.3) | $4.2 \times 10^{-5}$ | 8.1 (3.0 - 22.2) | $8.1 \times 10^{-3}$ | 0.9 | 0 | Other[126,127] |
| *VPS53* | $8.0 \times 10^{-5}$ | 6.1 (2.5 - 14.8) | $1.1 \times 10^{-5}$ | 6.9 (2.9 - 16.2) | $2.6 \times 10^{-3}$ | 1.0 | 0 | Other[128] |
| *ARHGAP1* | $9.4 \times 10^{-5}$ | 9.6 (3.1 - 29.7) | $9.4 \times 10^{-5}$ | 9.6 (3.1 - 29.7) | $2.9 \times 10^{-3}$ | 1.0 | 0 | Other[129,130] |
| *MRAP2* | $9.9 \times 10^{-5}$ | 17.1 (4.1 - 71.4) | $8.4 \times 10^{-6}$ | 20.7 (5.5 - 78.5) | $3.5 \times 10^{-3}$ | 1.0 | 0 | - |

Q, Cochran's Q statistic.
$I^2$, percentage of variation in effect estimates attributable to between-study heterogeneity rather than chance.

Supplementary Table 3. Results of the analysis of the combination of rare protein truncating variants (PTVs) and rare splice-altering variants in genes associated with endometrial cancer at exome-wide significance ($p < 2.5 \times 10^{-6}$) in the meta-analysis of UK Biobank (UKB) and 100,000 Genomes (100KG) Project datasets using logistic regression. The table shows predicted deleterious MSV carrier counts among cases and controls, together with relative risks (RRs) and 95% confidence intervals (CIs) from the UKB prevalent and incident meta-analyses, the 100KG analysis, and the meta-analyses combining UKB and 100KG. The asterisk (*) signifies genes that have previously been established as associated with endometrial cancer risk, and the diamond (◊) signifies genes that showed strong association only when splice-altering variants as predicted by SpliceAI were added to the analysis.

| Gene | UKB Counts (Prevalent Cases/ Controls) | UKB Counts (Incident Cases/ Controls) | UKB Meta Analysis (Prevalent and Incident Cohorts) | | 100K Genomes (Cases/ Controls) | 100KG Analysis | | Meta-analysis (UKB and 100KG) | |
|---|---|---|---|---|---|---|---|---|---|
| | | | Relative Risk (95% CI) | p value | | Odds Ratio (95% CI) | p value | Relative Risk (95% CI) | p value ($< 2.5\times10^{-6}$) |
| *MSH6** | 17/292 | 22/292 | 10.2 (7.4 - 14.1) | $1.2\times10^{-45}$ | 7/5 | 25.3 (7.9 - 81.0) | $5.1\times10^{-8}$ | 10.9 (8.0 - 14.8) | $1.3\times10^{-51}$ |
| *MLH1** | 4/14 | 2/14 | 35.8 (15.0 - 85.6) | $8.1\times10^{-16}$ | 1/5 | 3.3 (0.4 - 28.0) | 0.28 | 25.6 (11.4 - 57.3) | $3.6\times10^{-15}$ |
| *MSH2** | 5/57 | 1/57 | 12.5 (5.4 - 28.7) | $2.7\times10^{-9}$ | 3/6 | 8.4 (2.1 - 33.9) | $2.8\times10^{-3}$ | 11.3 (5.5 - 23.0) | $3.1\times10^{-11}$ |
| *BRWD1*◊ | 0/92 | 7/92 | 9.2 (4.4 - 19.4) | $4.6\times10^{-9}$ | ¼ | 4.5 (0.5 - 40.3) | 0.18 | 8.6 (4.2 - 17.3) | $2.2\times10^{-9}$ |
| *PMAIP1* | 0/15 | 3/15 | 22.5 (7.2 - 69.8) | $7.4\times10^{-8}$ | 0/4 | - | - | 22.5 (7.2 - 69.8) | $7.4\times10^{-8}$ |
| *UBE2V2* | 0/5 | 2/5 | 40.1 (10.0 - 160.9) | $1.9\times10^{-7}$ | 0/0 | - | - | 40.1 (10.0 - 160.9) | $1.9\times10^{-7}$ |
| *PHKG2* | 1/23 | 3/23 | 13.1 (4.9 - 35.0) | $3.3\times10^{-7}$ | 0/12 | - | 0.94 | 13.1 (4.9 - 35.0) | $3.3\times10^{-7}$ |
| *ERMN*◊ | 0/6 | 2/6 | 36.2 (9.0 - 145.1) | $4.0\times10^{-7}$ | 0/1 | - | 0.96 | 36.2 (9.0 - 145.0) | $4.0\times10^{-7}$ |
| *INTS15* | 1/3 | 1/3 | 46.3 (10.5 - 203.9) | $4.1\times10^{-7}$ | 0/0 | - | - | 46.3 (10.5 - 203.9) | $4.1\times10^{-7}$ |
| *RP1*◊ | 0/93 | 6/93 | 7.7 (3.4 - 17.2) | $6.3\times10^{-7}$ | 0/0 | - | - | 7.7 (3.4 - 17.2) | $6.3\times10^{-7}$ |
| *RHEBL1* | 1/12 | 2/12 | 18.4 (5.8 - 58.2) | $6.9\times10^{-7}$ | 0/1 | - | 0.97 | 18.4 (5.8 - 58.2) | $6.9\times10^{-7}$ |

| | | | | | | | | | |
|---|---|---|---|---|---|---|---|---|---|
| *ACRV1* | 1/26 | 3/26 | 11.9 (4.4 - 32.0) | $8.5\times10^{-7}$ | 0/0 | - | - | 11.9 (4.4 - 32.0) | $8.5x10^{-7}$ |
| *FNDC1*◊ | 3/131 | 6/131 | 5.2 (2.7 - 10.0) | $9.4\times10^{-7}$ | 0/11 | - | 0.95 | 5.2 (2.7 - 10.0) | $9.5x10^{-7}$ |
| *PID1* | 0/7 | 2/7 | 32.1 (8.0 - 128.8) | $1.0\times10^{-6}$ | 0/0 | - | - | 32.1 (8.0 - 128.8) | $1.0x10^{-6}$ |
| *SUPT16H*◊ | 0/7 | 2/7 | 31.2 (7.8 - 124.9) | $1.2\times10^{-6}$ | 0/0 | - | - | 31.2 (7.8 - 124.9) | $1.2x10^{-6}$ |
| *FGF4* | 0/7 | 2/7 | 31.0 (7.7 - 124.0) | $1.3\times10^{-6}$ | 0/0 | - | - | 31.0 (7.7 - 124.0) | $1.3x10^{-6}$ |
| *URAD*◊ | 2/9 | 0/9 | 42.0 (9.0 - 195.7) | $1.9\times10^{-6}$ | 0/2 | - | 0.97 | 42.0 (9.0 - 195.7) | $1.9x10^{-6}$ |
| *STK32C* | 0/22 | 3/22 | 15.7 (5.1 - 48.8) | $1.9\times10^{-6}$ | 0/1 | - | 0.96 | 15.7 (5.1 - 48.8) | $1.9x10^{-6}$ |

Supplementary Table 4. Genes with rare predicted deleterious missense variants (MSVs) associated with $p < 10^{-4}$ in the meta-analysis combining whole-exome sequencing data from the UK Biobank (UKB) and whole-genome sequencing data from the 100,000 Genomes Project (100KG). Results are presented for meta-analyses using standard logistic regression, Firth's bias-reduced regression and SKAT-O test. Heterogeneity between UKB and 100KG was assessed using Cochran's Q statistic and the $I^2$ statistic. Previous evidence of associations with endometrial, colorectal or other cancers is also reported.

| **Gene** | **Logistic regression meta-analysis** | | **Firth's regression meta-analysis** | | **SKAT-O meta-analysis** | **Heterogeneity** | | | **Previous cancer evidence** |
|---|---|---|---|---|---|---|---|---|---|
| | **p value** | **Relative Risk (95% CI)** | **Firth's p value** | **p value** | **p value** | **Q statistic** | **p value of Q statistic** | **$I^2$** | |
| *GPC5* | $3.0x10^{-8}$ | 6.5 (3.3 - 12.5) | $6.0x10^{-9}$ | 6.6 (3.5 - 12.4) | $2.03x10^{-9}$ | 1.1 | 0.3 | 0.09 | Other[131,132] |
| *PXN* | $9.6x10^{-8}$ | 11.9 (4.8 - 29.6) | $6.6x10^{-9}$ | 13.3 (5.6 - 32.0) | $1.39x10^{-16}$ | $4.8x10^{-3}$ | 0.9 | 0 | Colorectal[133–135] |
| *ARGLU1* | $1.6x10^{-7}$ | 17.7 (6.0 - 51.6) | $5.9x10^{-9}$ | 20.5 (7.4 - 56.8) | $9.17x10^{-7}$ | $4.8x10^{-1}$ | 0.5 | 0 | Other[136,137] |
| *MIER3* | $4.6x10^{-7}$ | 13.7 (5.0 - 37.9) | $3.4x10^{-8}$ | 15.2 (5.8 - 40.1) | $4.96x10^{-4}$ | $4.4x10^{-3}$ | 0.9 | 0 | Other[138,139] |
| *COA1* | $6.5x10^{-7}$ | 18.5 (5.9 - 58.2) | $4.4x10^{-8}$ | 21.0 (7.1 - 62.4) | - | 0.0 | - | - | Colorectal[140–142] |
| *PTEN* | $6.5x10^{-7}$ | 9.7 (4.0 - 23.7) | $3.6x10^{-8}$ | 10.7 (4.6 - 24.8) | $7.35x10^{-23}$ | 1.6 | 0.2 | 0.4 | Endometrial[143–145] |
| *LIMCH1* | $1.2x10^{-6}$ | 17.1 (5.4 - 53.5) | $1.2x10^{-6}$ | 17.1 (5.4 - 53.5) | - | 0.0 | - | - | Other[146–148] |
| *PCIF1* | $1.2x10^{-6}$ | 5.7 (2.8 - 11.4) | $2.1x10^{-7}$ | 6.1 (3.1 - 12.0) | $3.48x10^{-16}$ | $6.5x10^{-3}$ | 0.9 | 0 | Colorectal[149–151] |
| *ASB9* | $1.7x10^{-6}$ | 41.3 (9.0 - 189.6) | $1.2x10^{-7}$ | 48.9 (11.6 - 206.6) | - | 0.0 | - | - | Colorectal[152] |
| *SCD* | $3.7x10^{-6}$ | 7.3 (3.1 - 17.0) | $5.4x10^{-7}$ | 8.0 (3.5 - 17.9) | - | $2.0x10^{-1}$ | 0.7 | 0 | Other[153,154] |
| *RBFA* | $7.0x10^{-6}$ | 9.5 (3.6 - 25.4) | $6.9x10^{-6}$ | 9.5 (3.6 - 25.4) | $9.03x10^{-5}$ | $5.5x10^{-3}$ | 0.9 | 0 | - |
| *TTR* | $8.2x10^{-6}$ | 26.6 (6.3 - 112.4) | $6.3x10^{-7}$ | 32.2 (8.2 - 126.2) | - | 0.0 | - | - | Other[155,156] |
| *CDK11B* | $8.6x10^{-6}$ | 8.5 (3.3 - 21.8) | $3.7x10^{-7}$ | 10.2 (4.2 - 24.9) | $1.73x10^{-7}$ | $3.4x10^{-1}$ | 0.6 | 0 | Other[157] |
| *PRAMEF15* | $9.8x10^{-6}$ | 6.9 (2.9 - 16.2) | $5.0x10^{-6}$ | 7.2 (3.1 - 16.7) | $1.13x10^{-4}$ | $3.8x10^{-1}$ | 0.5 | 0 | Other[158,159] |
| *CACNG8* | $9.9x10^{-6}$ | 4.8 (2.4 - 9.6) | $3.4x10^{-6}$ | 4.9 (2.5 - 9.7) | - | $6.7x10^{-1}$ | 0.4 | 0 | - |
| *SYCE1L* | $1.0x10^{-5}$ | 22.9 (5.7 - 91.8) | $1.0x10^{-5}$ | 22.9 (5.7 - 91.8) | - | $3.6x10^{-3}$ | 1.0 | 0 | - |

| | | | | | | | | | |
|---|---|---|---|---|---|---|---|---|---|
| *FAM43B* | 1.2x10$^{-5}$ | 4.9 (2.4 - 9.9) | 8.8x10$^{-6}$ | 5.0 (2.5 - 10.1) | 1.10x10$^{-2}$ | 1.2 | 0.3 | 0.2 | Colorectal[160], Other[161] |
| *RMI2* | 1.3x10$^{-5}$ | 3.8 (2.1 - 7.0) | 7.0x10$^{-6}$ | 3.9 (2.2 - 7.2) | 1.39x10$^{-1}$ | 1.4x10$^{-3}$ | 1.0 | 0 | Other[162–164] |
| *ITPRID2* | 1.5x10$^{-5}$ | 2.8 (1.8 - 4.5) | 3.3x10$^{-6}$ | 3.0 (1.9 - 4.7) | - | 1.4 | 0.2 | 0.3 | Other[165] |
| *THEMIS* | 1.6x10$^{-5}$ | 4.4 (2.2 - 8.6) | 3.3x10$^{-6}$ | 4.7 (2.4 - 9.0) | 1.98x10$^{-3}$ | 2.2 | 0.1 | 0.5 | - |
| *BICRAL* | 1.7x10$^{-5}$ | 8.6 (3.2 - 23.0) | 1.8x10$^{-5}$ | 8.6 (3.2 - 23.0) | 8.21x10$^{-6}$ | 6.4x10$^{-1}$ | 0.4 | 0 | - |
| *THAP2* | 1.8x10$^{-5}$ | 6.1 (2.7 - 14.0) | 2.8x10$^{-6}$ | 6.7 (3.0 - 14.8) | 1.90x10$^{-7}$ | 1.2 | 0.3 | 0.1 | - |
| *CNPY3* | 1.8x10$^{-5}$ | 7.1 (2.9 - 17.5) | 2.2x10$^{-6}$ | 8.0 (3.4 - 19.0) | 5.68x10$^{-6}$ | 3.4x10$^{-3}$ | 1.0 | 0 | Colorectal[166] |
| *PRL* | 2.0x10$^{-5}$ | 9.3 (3.3 - 26.0) | 1.4x10$^{-5}$ | 9.6 (3.5 - 26.7) | - | 7.8x10$^{-3}$ | 0.9 | 0 | Other[167,168] |
| *ALG1L2* | 2.0x10$^{-5}$ | 20.4 (5.1 - 81.8) | 2.0x10$^{-5}$ | 20.4 (5.1 - 81.8) | - | 3.7x10$^{-3}$ | 1.0 | 0 | - |
| *PIP4K2A* | 2.2x10$^{-5}$ | 5.7 (2.5 - 12.7) | 8.2x10$^{-6}$ | 5.8 (2.7 - 12.6) | 3.18x10$^{-6}$ | 4.9x10$^{-3}$ | 0.9 | 0 | Other[169] |
| *MTFR2* | 2.5x10$^{-5}$ | 23.5 (5.4 - 101.7) | 1.4x10$^{-6}$ | 28.5 (7.3 - 110.7) | - | 4.8x10$^{-3}$ | 0.9 | 0 | Endometrial[170], Other[171,172] |
| *UBXN7* | 2.6x10$^{-5}$ | 12.3 (3.8 - 39.6) | 2.3x10$^{-6}$ | 14.1 (4.7 - 42.4) | 5.69x10$^{-3}$ | 4.3x10$^{-3}$ | 0.9 | 0 | Other[173,174] |
| *SS18L1* | 2.8x10$^{-5}$ | 12.1 (3.8 - 39.0) | 2.3x10$^{-6}$ | 14.0 (4.7 - 41.8) | - | 3.8x10$^{-3}$ | 1.0 | 0 | Other[175,176] |
| *CLTRN* | 2.9x10$^{-5}$ | 19.3 (4.8 - 77.4) | 2.9x10$^{-5}$ | 19.3 (4.8 - 77.4) | 1.78x10$^{-5}$ | 0.0 | - | - | Other[177] |
| *FBXO31* | 3.5x10$^{-5}$ | 4.3 (2.2 - 8.7) | 1.2x10$^{-5}$ | 4.5 (2.3 - 8.9) | 2.58x10$^{-5}$ | 3.6x10$^{-3}$ | 1.0 | 0 | Endometrial[178], Other[179] |
| *CLINT1* | 4.8x10$^{-5}$ | 4.4 (2.1 - 8.9) | 5.3x10$^{-6}$ | 4.9 (2.5 - 9.7) | 1.44x10$^{-8}$ | 3.6x10$^{-1}$ | 0.6 | 0 | Colorectal[180,181] |
| *BARX1* | 5.2x10$^{-5}$ | 3.0 (1.8 - 5.0) | 1.1x10$^{-5}$ | 3.2 (1.9 - 5.3) | 5.15x10$^{-4}$ | 3.7x10$^{-1}$ | 0.5 | 0 | Endometrial[182], Colorectal[183], Other[184,185] |
| *AKR1A1* | 5.5x10$^{-5}$ | 5.4 (2.4 - 12.1) | 1.2x10$^{-5}$ | 5.8 (2.6 - 12.7) | 1.34x10$^{-2}$ | 4.9x10$^{-3}$ | 0.9 | 0 | - |
| *GALNT7* | 5.7x10$^{-5}$ | 4.0 (2.0 - 7.9) | 8.6x10$^{-6}$ | 4.4 (2.3 - 8.6) | 2.93x10$^{-5}$ | 9.2x10$^{-2}$ | 0.8 | 0 | Colorectal[186,187], Other[188,189] |
| *HMGA2* | 6.0x10$^{-5}$ | 17.1 (4.3 - 68.7) | 6.0x10$^{-5}$ | 17.1 (4.3 - 68.7) | - | 0.0 | - | - | Endometrial[190,191] |
| *ZNF418* | 6.1x10$^{-5}$ | 3.7 (2.0 - 7.1) | 1.0x10$^{-5}$ | 4.1 (2.2 - 7.6) | - | 1.8x10$^{-1}$ | 0.7 | 0 | - |
| *GNAi2* | 6.8x10$^{-5}$ | 6.0 (2.5 - 14.4) | 1.9x10$^{-5}$ | 6.3 (2.7 - 14.6) | 3.28x10$^{-4}$ | 1.2 | 0.3 | 0.2 | Other[192,193] |
| *KCNK2* | 6.9x10$^{-5}$ | 19.3 (4.5 - 83.0) | 4.4x10$^{-6}$ | 23.3 (6.1 - 89.4) | - | 4.2x10$^{-3}$ | 0.9 | 0 | Other[194,195] |
| *FYTTD1* | 7.3x10$^{-5}$ | 5.7 (2.4 - 13.3) | 8.9x10$^{-6}$ | 6.4 (2.8 - 14.5) | - | 2.1x10$^{-1}$ | 0.6 | 0 | - |

| | | | | | | | | | |
|---|---|---|---|---|---|---|---|---|---|
| *H2BC15* | $7.4x10^{-5}$ | 4.6 (2.2 - 9.7) | $1.7x10^{-5}$ | 4.8 (2.3 - 9.8) | $8.43x10^{-4}$ | $2.8x10^{-1}$ | 0.6 | 0 | - |
| *SHFL* | $7.7x10^{-5}$ | 16.4 (4.1 - 65.7) | $7.7x10^{-5}$ | 16.4 (4.1 - 65.8) | $1.15x10^{-5}$ | $6.3x10^{-3}$ | 0.9 | 0 | - |
| *JRKL* | $8.3x10^{-5}$ | 5.9 (2.4 - 14.1) | $2.4x10^{-5}$ | 6.1 (2.6 - 14.2) | - | $3.8x10^{-3}$ | 1.0 | 0 | - |
| *GMPPA* | $8.6x10^{-5}$ | 2.8 (1.7 - 4.6) | $3.0x10^{-5}$ | 2.9 (1.7 - 4.7) | - | 1.7 | 0.2 | 0.4 | Other[196,197] |

Q, Cochran's Q statistic.
$I^2$, percentage of variation in effect estimates attributable to between-[study] heterogeneity rather than chance.

Supplementary Table 5. Results for genes with previously reported evidence of association with endometrial cancer, analysed using logistic regression. Carrier counts among cases and controls, relative risks (RRs) and 95% confidence intervals (CIs) are presented for rare protein-truncating variants (PTVs) and rare predicted deleterious missense variants (MSVs). Results are shown separately for the UKB prevalent and incident meta-analyses, the 100KG analysis, and the meta-analyses combining UKB and 100KG.

| **Gene** | **UKB Counts (Prevalent Cases/ Controls)** | **UKB Counts (Incident Cases/ Controls)** | **UKB Meta Analysis (Prevalent and Incident Cohorts)** | | **100K Genomes (Cases/ Controls)** | **100KG Analysis** | | **Meta-analysis (UKB and 100KG)** | |
|---|---|---|---|---|---|---|---|---|---|
| | | | **Relative Risk (95% CI)** | **p value** | | **Odds Ratio (95% CI)** | **p value** | **Relative Risk (95% CI)** | **p value** |
| **Protein-truncating variants (PTVs)** | | | | | | | | | |
| *MSH6* | 16/63 | 17/63 | 38.0 (26.4 - 54.5) | $5.0\times10^{-86}$ | 7/5 | 25.3 (7.9 - 81.0) | $5.1\times10^{-8}$ | 36.6 (25.9 - 51.8) | $2.2\text{x}10^{-92}$ |
| *MLH1* | 4/9 | 2/9 | 52.5 (21.3 - 129.4) | $8.1\times10^{-18}$ | 0/1 | - | 0.96 | 52.5 (21.3 - 129.4) | $8.1\text{x}10^{-18}$ |
| *MSH2* | 4/38 | 1/38 | 14.4 (5.8 - 36.0) | $1.1\times10^{-8}$ | 3/5 | 10.0 (2.4 - 42.4) | $1.7\times10^{-3}$ | 13.0 (6.0 - 28.2) | $7.6\text{x}10^{-11}$ |
| *POLE* | 5/261 | 2/261 | 2.5 (1.2 - 5.3) | $1.5\times10^{-2}$ | 2/14 | 2.4 (0.5 - 10.6) | 0.25 | 2.5 (1.3 - 4.9) | $7.3\text{x}10^{-3}$ |
| *POLD1* | 0/89 | 1/89 | 1.4 (0.2 - 9.8) | 0.75 | 0/10 | - | 0.95 | 1.4 (0.2 – 9.8) | $7.5\text{x}10^{-1}$ |
| *PTEN* | 0/2 | 0/2 | - | - | 2/0 | $5.95\text{x}10^{-6}$ ($3.13\text{x}10^{-189}$– $1.13\text{x}10^{202}$ | 0.95 | $6.0\text{x}10^{+6}$ (0.0 - $1.1\text{x}10^{+202}$) | $6.8\text{x}10^{-2}$ |
| *PMS2* | 1/77 | 1/77 | 2.0 (0.5 - 8.2) | 0.32 | 2/4 | 9.2 (1.7 - 50.8) | $1.1\times10^{-2}$ | 3.7 (1.3 - 10.9) | $1.7\text{x}10^{-2}$ |
| *EPCAM* | 0/29 | 0/29 | - | 0.95 | 0/2 | - | 0.97 | $7.8\text{x}10^{-5}$ (0.0 - $2.3\text{x}10^{+105}$) | $9.4\text{x}10^{-1}$ |
| *BRCA1* | 2/85 | 2/85 | 3.6 (1.4 - 9.7) | $1.1\times10^{-2}$ | 0/6 | - | 0.96 | 3.6 (1.4 - 9.7) | $1.1\text{x}10^{-2}$ |
| *BRCA2* | 5/262 | 6/262 | 3.2 (1.8 - 5.8) | $1.3\times10^{-4}$ | 0/6 | - | 0.96 | 3.2 (1.8 - 5.8) | $1.3\text{x}10^{-4}$ |
| **Missense variants (MSVs) predicted to be deleterious** | | | | | | | | | |
| *PTEN* | 1/24 | 3/24 | 12.7 (4.7 - 33.9) | $4.6\times10^{-7}$ | 1/6 | 2.8 (0.3 - 23.5) | 0.34 | 9.7 (4.0 - 23.7) | $6.5\text{x}10^{-7}$ |
| *MSH6* | 16/3396 | 45/3396 | 1.4 (1.1 - 1.8) | $9.3\times10^{-3}$ | 8/25 | 5.2 (2.3 - 11.6) | $5.6\times10^{-5}$ | 1.6 (1.2 - 2.0) | $2.1\text{x}10^{-4}$ |

| | | | | | | | | | |
|---|---|---|---|---|---|---|---|---|---|
| *MLH1* | 15/2253 | 22/2253 | 1.2 (0.9 - 1.7) | 0.18 | 5/52 | 1.6 (0.6 - 4.0) | 0.33 | 1.3 (0.9 - 1.7) | $1.2x10^{-1}$ |
| *MSH2* | 5/829 | 10/829 | 1.4 (0.8 - 2.3) | 0.21 | 4/63 | 1.0 (0.4 - 2.8) | 0.97 | 1.3 (0.8 - 2.1) | $2.5x10^{-1}$ |
| *POLE* | 20/6532 | 54/6531 | 0.9 (0.7 - 1.1) | 0.30 | 6/81 | 1.2 (0.5 - 2.8) | 0.65 | 0.9 (0.7 - 1.1) | $3.8x10^{-1}$ |
| *POLD1* | 2/631 | 5/631 | 0.9 (0.4 - 1.8) | 0.68 | 3/51 | 1.0 (0.3 - 3.1) | 0.94 | 0.9 (0.5 - 1.7) | $7.0x10^{-1}$ |
| *PMS2* | 97/18370 | 139/18364 | 1.0 (0.9 - 1.1) | 0.71 | 1/31 | 0.5 (0.1 - 4.0) | 0.55 | 1.0 (0.8 - 1.1) | $6.9x10^{-1}$ |
| *EPCAM* | 0/93 | 1/93 | 1.3 (0.2 - 9.4) | 0.78 | 1/8 | 1.9 (0.2 - 15.2) | 0.55 | 1.6 (0.4 - 6.5) | $5.4x10^{-1}$ |
| *BRCA1* | 12/2362 | 24/2362 | 1.2 (0.8 - 1.6) | 0.36 | 2/6 | 5.9 (1.2 - 29.4) | $3.1\times10^{-2}$ | 1.2 (0.9 - 1.7) | $1.8x10^{-1}$ |
| *BRCA2* | 2/880 | 11/880 | 1.3 (0.7 - 2.2) | 0.38 | 3/69 | 0.7 (0.2 - 2.3) | 0.58 | 1.1 (0.7 - 1.9) | $5.8x10^{-1}$ |

Supplementary Table 6. The number of selected variants within the known endometrial cancer genes in each category before and after filtering based on ClinVar annotations.

| Dataset | Gene | Number of selected protein-truncating variants | | Number of selected predicted deleterious missense variants | | Number of selected splice-altering variants | |
|---|---|---|---|---|---|---|---|
| | | Before filtering | After filtering | Before filtering | After filtering | Before filtering | After filtering |
| UK Biobank | *MSH2* | 40 | 31 | 259 | 8 | 9 | 1 |
| | *MSH6* | 150 | 112 | 299 | 10 | 4 | 0 |
| | *MLH1* | 33 | 38 | 139 | 8 | 2 | 0 |
| | *PMS2* | 82 | 45 | 129 | 83 | 3 | 1 |
| | *PTEN* | 15 | 6 | 56 | 13 | 1 | 0 |
| 100K Genomes | *MSH2* | 5 | 4 | 26 | 0 | 1 | 0 |
| | *MSH6* | 8 | 6 | 27 | 3 | 0 | 0 |
| | *MLH1* | 1 | 1 | 30 | 5 | 3 | 0 |
| | *PMS2* | 9 | 6 | 21 | 8 | 0 | 0 |
| | *PTEN* | 3 | 3 | 6 | 1 | 0 | 0 |

Supplementary Figure 1. Flow chart illustrating the participant selection from the UK Biobank.

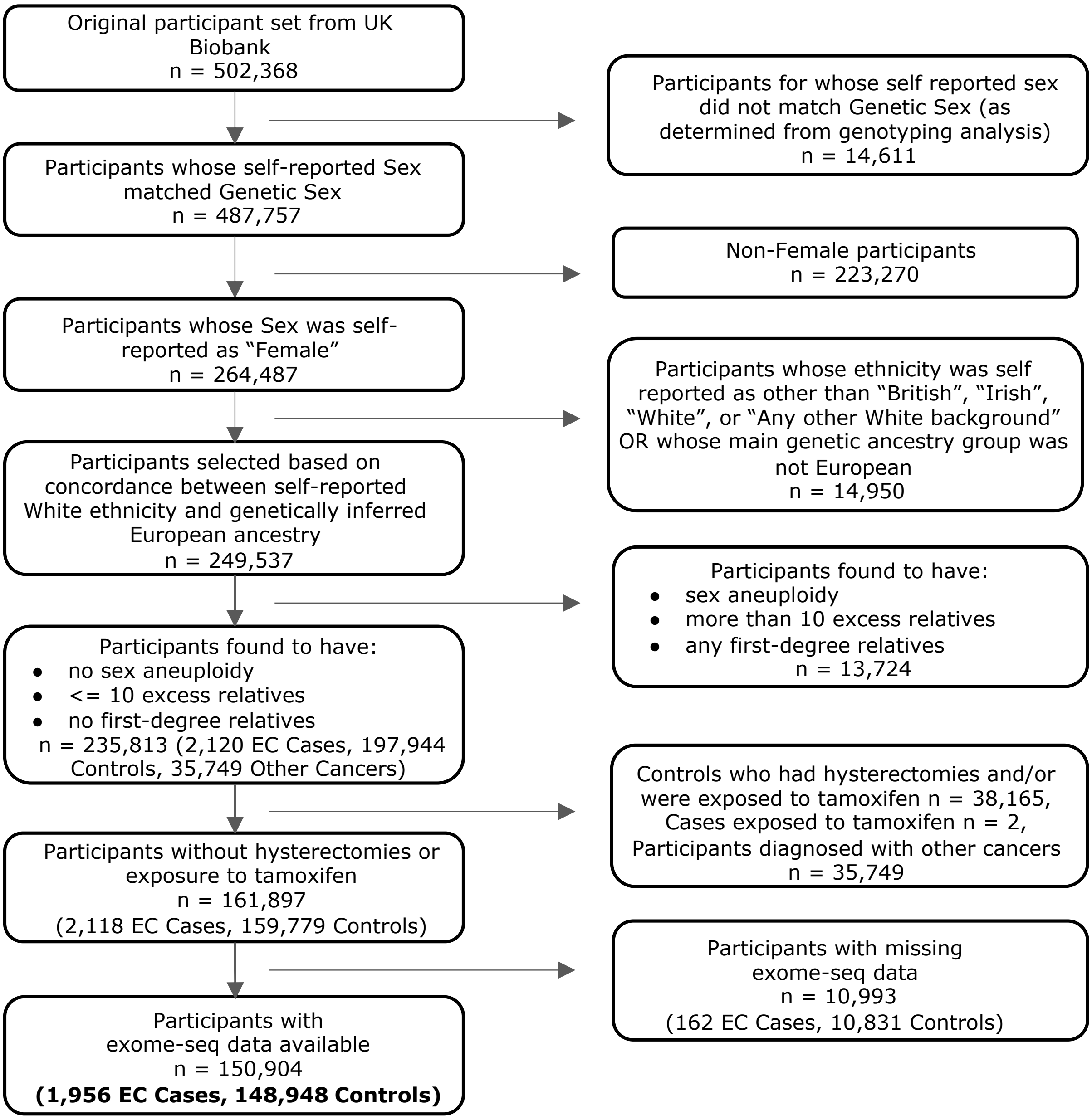

Supplementary Figure 2. Flow chart illustrating participant selection for a) cases and b) controls from the 100,000 Genomes project.

a) CASES

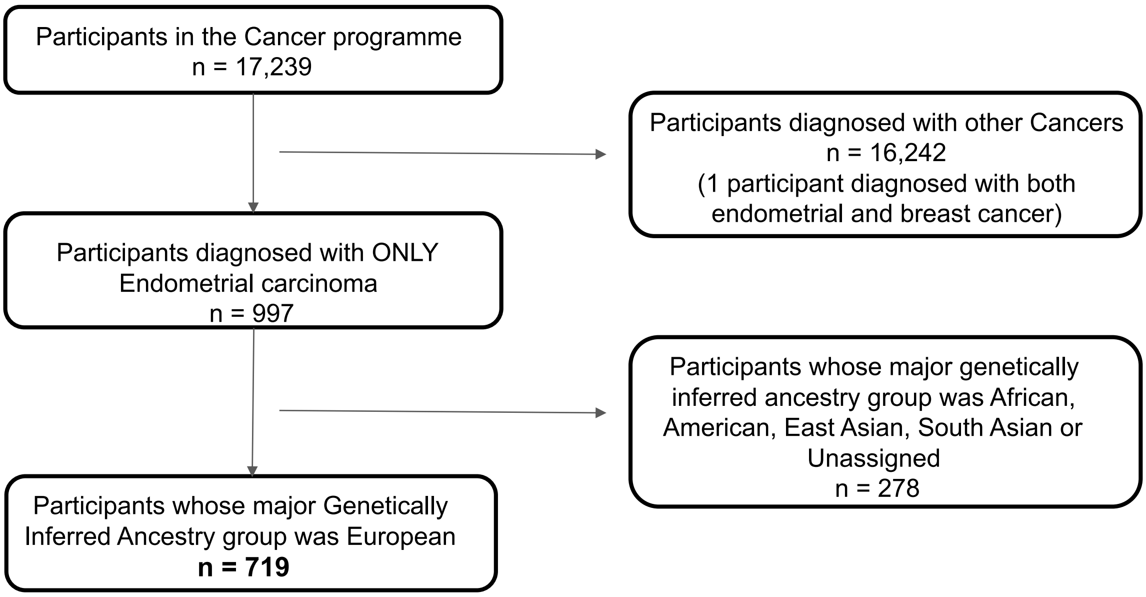


b) CONTROLS

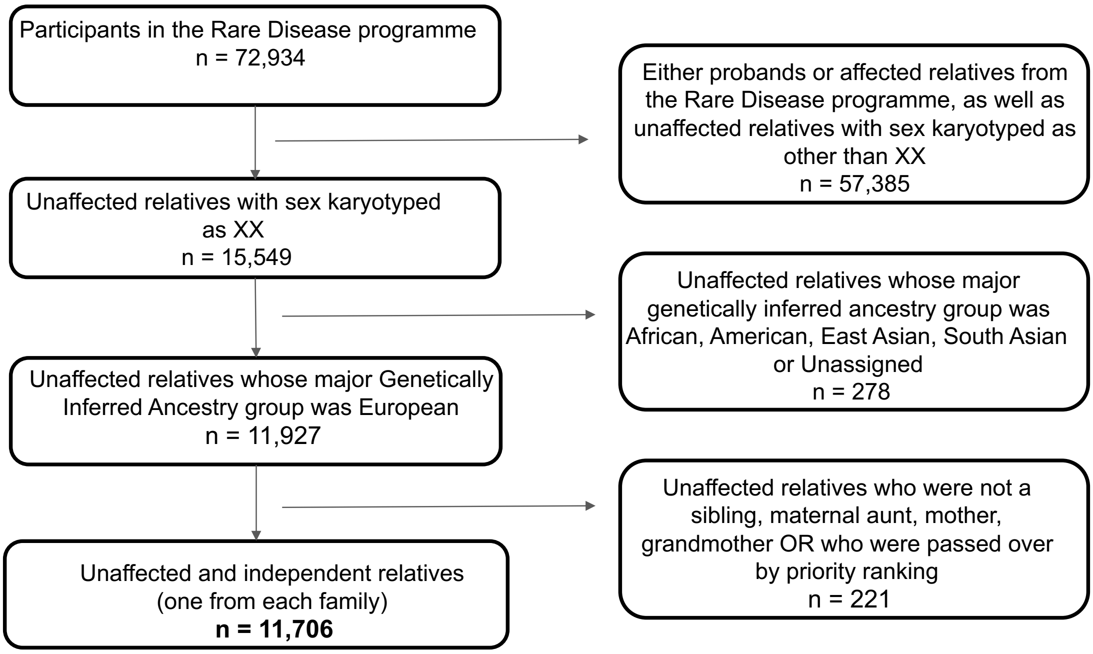
